# LOW LEVELS OF DIRECT ORAL ANTICOAGULANTS AT STEADY STATE IN ATRIAL FIBRILLATION PATIENTS WHO LATER DEVELOP THROMBOTIC COMPLICATIONS; THE PROSPECTIVE MAS (MEASURE AND SEE) STUDY

**DOI:** 10.1101/2023.12.08.23299746

**Authors:** Sophie Testa, Gualtiero Palareti, Cristina Legnani, Claudia Dellanoce, Michela Cini, Oriana Paoletti, Antonio Ciampa, Emilia Antonucci, Daniela Poli, Rossella Morandini, Maurizio Tala, Paolo Chiarugi, Rita Carlotta Santoro, Angela Maria Iannone, Erica De Candia, Pasquale Pignatelli, Elena Maria Faioni, Antonio Chistolini, Maria del Pilar Esteban, Marco Marietta, Armando Tripodi, Alberto Tosetto, MAS Study group

**Affiliations:** Centro Emostasi e Trombosi, UO Laboratorio Analisi Chimico-Cliniche e Microbiologiche, ASST Cremona, Cremona, Italy; Fondazione Arianna Anticoagulazione, Bologna, Italy; Centro Emostasi, UOC Laboratorio Analisi, Ospedale S.G. Moscati, Avellino, Italy; Malattie Aterotrombotiche, AOU Careggi, Firenze, Italy; UO di Analisi chimico cliniche, Azienda Ospedaliero Universitaria Pisana, Pisa, Italy; Centro Emostasi e Trombosi, UO Emofilia e Patologie della Coagulazione, Dipartimento di Ematologia, Oncologia e Medicina Trasfusionale, Azienda Ospedaliero Universitaria Dulbecco, Catanzaro, Italy; UOSVD Sezione Trasfusionale, Ospedale Don Tonino Bello, Molfetta, Bari, Italy; UOSD Malattie Emorragiche e Trombotiche, Dipartimento Diagnostica per Immagini, Radioterapia Oncologica ed Ematologia, Fondazione Policlinico Universitario Agostino Gemelli IRCCS, Roma, Italy; UOC Medicina Interna e Prevenzione dell’Aterosclerosi, Dipartimento di Medicina Interna e Specialità Mediche, Azienda Ospedaliero-Universitaria Policlinico Umberto I, Roma, Italy; Servizio Immunologia e Medicina Trasfusionale, Ospedale San Paolo, ASST Santi Paolo e Carlo, Milano, Italy; UO Medicina Traslazionale e di Precisione, Dipartimento Medicina Interna e Specialità Mediche, Azienda Ospedaliero-Universitaria Policlinico Umberto I, Roma, Italy; UO Laboratorio Analisi, Dipartimento dei Servizi Diagnostici, Ospedale Oglio Po, ASST Cremona, Cremona, Italy; Struttura Complessa di Ematologia, Policlinico di Modena, Azienda Ospedaliera-Universitaria di Modena, Modena, Italy; Centro Emofila e Trombosi Angelo Bianchi Bonomi, presso la Fondazione IRCCS Ca’ Granda Ospedale Maggiore, Milano, Italy; UOC Ematologia, Centro Malattie Emorragiche e Trombotiche (CMET), AULSS 8 Berica Ospedale S. Bortolo, Vicenza, Italy

## Abstract

**Background:** Though effective and safe, treatment with direct oral anticoagulants (DOAC) in atrial fibrillation (AF) is still associated with thrombotic complications. Whether the measurement of DOAC levels may improve the efficacy of treatment is an open issue.

**Methods:** We carried out the observational, prospective, multicenter study [“Measure and See” Study (MAS) (NCT03803579)]. Blood was collected 15-30 days after starting DOAC treatment in AF patients, who were then followed for one year. Plasma samples were centralized, for DOAC levels measurement. Patients’ DOAC levels were converted into drug/dosage standardized values to allow a pooled analysis in a time-dependent, competitive-risk model. The measured values were transformed into standardized values by subtracting from the original values the overall mean value of each DOAC and divided by the standard deviation. Standardized values represent the distance of each value from the overall mean.

**Results:** Trough and peak DOAC levels were assessed in 1657 and 1303 patients, respectively. Twenty-one thrombotic complications were recorded during 1606 years of follow-up (incidence of 1.31% patient/years). 17/21 thrombotic events occurred in patients whose standardized activity levels were below the mean of each DOAC (zero); the incidence was the highest (4.08% patient/years) in patients whose standardized values were in the lowest class (below zero, ≤ - 1.00).

**Conclusions:** Early measurement of DOAC levels in AF patients allowed us to identify most of the subjects who, having low baseline DOAC levels, subsequently developed thrombotic complications. Further studies are warranted to assess whether thrombotic complications may be reduced by measuring baseline DOAC levels and modifying treatment when indicated.

## INTRODUCTION

Over the last years, clinical trials, metanalyses and clinical practice confirmed the efficacy and safety of direct oral anticoagulants (DOAC) for stroke prevention in patients with non-valvular atrial fibrillation (AF).^1–8^ Recent observational studies in AF patients showed that DOAC, compared to warfarin, had lower rates of stroke, systemic embolism, comparable rates of major bleedings (MB),^9^ and advantages in terms of risk reduction of intracranial bleeding and systemic embolism even in the elderly and frail populations.^10,11^ However, a non-negligible incidence of thrombotic and bleeding events has been recorded in clinical trials and in observational studies in patients receiving DOAC.^5,7,8^ Hence, the issue of improving the clinical management of DOAC-treated patients and further reducing the risk of complications during treatment is relevant.

DOAC are administered to AF patients based on patient characteristics such as age, comorbidity, body weight, and renal function, without dose adjustment based on DOAC concentration measurements. This is mainly based on results of pharmacokinetic studies, which indicated a predictable anticoagulant response and an effective prevention of excessive drug concentration.^12–16^ Furthermore, the registration trials were conducted at dose regimens adjusted for some patient characteristics or for concomitant use of associated interacting drugs, and not for measured DOAC levels.

Consequently, the measurement of DOAC levels has been recommended only in particular situations, such as bleeding or thrombotic complications, before urgent need of surgery or invasive procedures, use of antidotes, and also suggested in special patient populations, such as those with frailty, under or overweight, or treated for epilepsy.^17–19^ However, studies focusing on the measurement of plasma DOAC levels showed high inter-patient variability for all the DOAC and for all the doses used to treat patients.^20–24^

Moreover, post-marketing studies showed that the variability of DOAC levels was even higher than that reported in phase II and III studies.^25–29^ Indeed, observational studies reported that some of the patients treated at fixed doses, may have relatively high or low DOAC plasma levels, thus supporting the issue of assessing whether an early or periodical measurement might contribute to improving the quality of treatment and reducing risks of complications.^30,31^

In addition, very recent studies showed a relationship between low DOAC levels (generally measured after the events), the risk of ischemic stroke and its severity,^32^ and the risk of stroke recurrences.^33^ Finally, a pilot prospective multicenter study evidenced low plasma levels of DOAC in patients with AF, measured at the time of embolic stroke.^34^ Whether DOAC concentration measurement may be useful to better tailoring the dose and optimizing the risk-benefit of treatment remains an unsolved clinical problem.

The present study aimed to investigate whether low DOAC plasma levels, assessed at steady-state within the first month of treatment, are associated with thrombotic events during a one-year follow-up.

## MATERIAL AND METHODS

The Measure and See Study (MAS) (NCT03803579) is an observational, prospective cohort, multicenter study of patients with AF, who started treatment with one of the available DOAC (dabigatran, apixaban, edoxaban, and rivaroxaban) for therapy and prevention of thrombotic complications. The study was promoted and funded by the Arianna Anticoagulazione Foundation (Bologna, Italy) and conducted in Anticoagulation Clinics affiliated with the Italian Federation of Anticoagulation Centers (FCSA).

### Patient population

Consecutive patients with AF, without rheumatic mitral valve disease or mechanical heart valves, aged over 18 years, seen at the anticoagulation clinics from 27 August 2018 to 10 November 2022, who had started within one month anticoagulation with a DOAC, were enrolled in the study. Patients suitable for electrical cardioversion, or who have refused blood sampling, or did not accept follow-up for at least one year or who had other clinical indications for anticoagulant therapy were excluded from the study.

The choice of DOAC and dose used was left to the discretion of treating physicians. The study protocol included a recommendation for the participant centers to include patients, who were treated following the rules defined for each drug by the Italian regulatory agency (AIFA) and current clinical practice.

Each patient was given a unique anonymous identifying code to ensure anonymity, which was used to collect clinical information and identify biological samples. The following baseline characteristics were recorded in a specific electronic database: patient identification number, date of birth, gender, type of drug used and dose, weight, body mass index, kidney function [estimated by creatinine clearance (CrCl) according to the Cockroft-Gault formula], liver function (assessed by aspartate aminotransferase and alanine aminotransferase), diabetes, CHA2DS2-VASc score, previous stroke/transient ischemic attack (TIA), other comorbidities, concomitant medications, (with particular attention to antiplatelet drugs). Data were stored in the database located at a section of the Aruba cloud rented by the Arianna Anticoagulazione Foundation, which guaranteed storage, backup, and maintenance of the database.

The study was conducted in accordance with the ethical principles of the Declaration of Helsinki. Independent review board approval was obtained before all study-related activity from the Ethics Committee (EC) of the coordination center (Cremona) (approval number 14725; 02/05/2018) and from the ECs of all other centers. Written informed consent was obtained from each patient before enrolment. The study promoter provided measures to safeguard the subject’s privacy and the protection of personal data according to the EU GDPR 2016/679 and Italian law.

### Blood sampling and DOAC measurement

The study required a mandatory plasma collection for the measurement of the DOAC level for each enrolled patient. Venous blood sampling had to be performed at a steady state (within the first 2-4 weeks of initiation of treatment) and obtained at the time of trough (C-trough), immediately before the subsequent drug intake. It was up to the discretion of participating centers, depending on their availability and organization, to perform additional blood collection on the same day, 2 hours after the last intake (C-peak). Additional blood samples were used to perform ancillary laboratory tests (including blood cell count, CrCl, and liver enzymes). Plasma samples for DOAC measurement were collected in vacuum tubes (Vacutainer; Franklin Lakes, NJ, USA) containing 1/10 volumes of 3.2% trisodium citrate. Blood was centrifuged within one hour of collection at 2000x g for 20 min (controlled room temperature), and plasma samples were aliquoted in cryovials, identified locally to maintain patient’s anonymity, in volumes that allow for optimal DOAC testing ; sample vials were stored frozen (-80°C)^35^ at the participating centers and later centralized to the biobank of the Arianna Anticoagulazione Foundation (Bologna). Finally, aliquots were transferred (dry-ice and express courier) to the Haemostasis and Thrombosis Center of Cremona Hospital for DOAC measurement.

DOAC levels, expressed as drug concentration-equivalent (ng/mL), were measured by chromogenic assays using STA-ECA II (DiagnosticaStago, Asnieres-sur-Seine, France) for dabigatran, and STA-Liquid anti-Xa (DiagnosticaStago) for apixaban, edoxaban, and rivaroxaban (hemolyzed samples were discarded). Tests were calibrated using commercial plasmas with certified DOAC concentration supplied by the same manufacturer and performed on STA-R Max instrument (Diagnostica Stago). The results of DOAC levels for each patient were transmitted to the central database repository, and were not communicated to patients, participating centers, or attending physicians.

### Follow-up and outcomes

Participating centers were instructed to organize blood sampling and clinical follow-up as requested by the study. Follow-up, as defined by FCSA guidelines, included a clinical evaluation within the first month of treatment and a clinical check-up every 3 to 4 months for one year. All thromboembolic and bleeding complications, death and other events were recorded during the 12-month follow-up.

The predefined study outcomes were all thromboembolic complications, including objectively documented ischemic cerebral vascular events, systemic emboli, occurrence of acute venous thromboembolism (VTE), acute myocardial infarction (AMI), and thrombotic and cardiovascular deaths. An independent Adjudication Committee evaluated adverse events occurring during follow–up. The present report analyses of the data regarding the relationship between DOAC levels and the occurrence of thromboembolic complications during follow-up. The relationship between DOAC levels and bleeding events will be analyzed and reported separately.

### Statistical analysis

*Sample size.* Based on previous studies,^12,25^ we hypothesized that the annualized risk of the primary study outcome may be fourfold as high in the lowest quintile of DOAC distribution as compared to the highest. We assumed an annualized risk of primary study outcome of 1.25% patient/years (pt/y) and 5% pt/y in patients in the highest and lowest quintile of DOAC plasma concentration, respectively. Under this assumption, a total sample size of 1315 patients would be sufficient to refute a null hypothesis of equivalence with alpha and beta errors of 0.05 and 0.2, respectively.

*Analysis plan.* Since the absolute C-trough and C-peak DOAC plasma concentration are drug and dose dependent, we standardized each measured absolute value using their drug- and dose-specific mean and standard deviation. Standardized values represent the distance of each value from the drug mean distribution and may be therefore pooled to evaluate the effect of drug levels, irrespective of the DOAC type and administration (OID or BID). Supplemental Figure 1 shows the correlation between unstandardized (absolute) and standardized plasma drug concentrations.

The outcome incidence rates were computed for all patients with at least one measured plasma DOAC concentration (C-trough or C-peak). Patients were censored at the end of the study or after the occurrence of a qualifying event. For the primary analysis, we used a Cox regression model allowing for competing risk according to Fine & Gray^36^ to model the occurrence of the primary study outcomes. The regression model included as a possible confounder CHA2DS2VASc score, body mass index, and the CrCl and concomitant antiplatelet use. Deaths occurring for causes other than thromboembolic events were considered as competing events. Akaike information criteria (AIC) was used to evaluate the goodness of fit of the Fine & Gray model.

As an exploratory analysis, we evaluated the incidence of the primary thrombotic outcome stratified by standardized values. For this analysis, Kaplan-Meier survival curves were plotted to estimate the cumulative incidence of thrombotic outcomes in patients with standardized values in the lowest class compared to those in the higher classes, and HR and their 95% CI were calculated. The variable CHA2DS2VASc score (≥4 vs <4) was arbitrarily categorized according to the median values as cut-off. Data were analyzed with the use of Prism software (Version 9.3.1, GraphPad Software Incorporated, San Diego, CA) and SPSS software (version 11.0 SPSS Inc., IBM, Armonk, NY), and R (version 4.3.1, R Foundation for Statistical Computing, Vienna). Raw data and scripts used for analysis are available upon request to the Authors at osf.io (https://osf.io, Center for Open Science, Charlottesville, VA).

## RESULTS

### Characteristics of patient population

The flow chart of the study population is shown in Figure 1. A total of 1718 patients, who started a DOAC treatment for non-valvular AF, were included in the study. After the exclusions (detailed in Figure 1), 1657 patients had blood sampling for DOAC levels measurement 15 to 30 days from the start of treatment (steady state). The clinical history for one year follow-up was collected in 1345 patients and for a shorter period for the remaining patients: 139 because the study was stopped; 173 patients were censored (72 for occurrence of thrombotic or bleeding complications, 37 for death, 64 for other reasons). The main demographic and clinical characteristics of the 1657 investigated patients are shown in Table 1.

**Figure 1.**
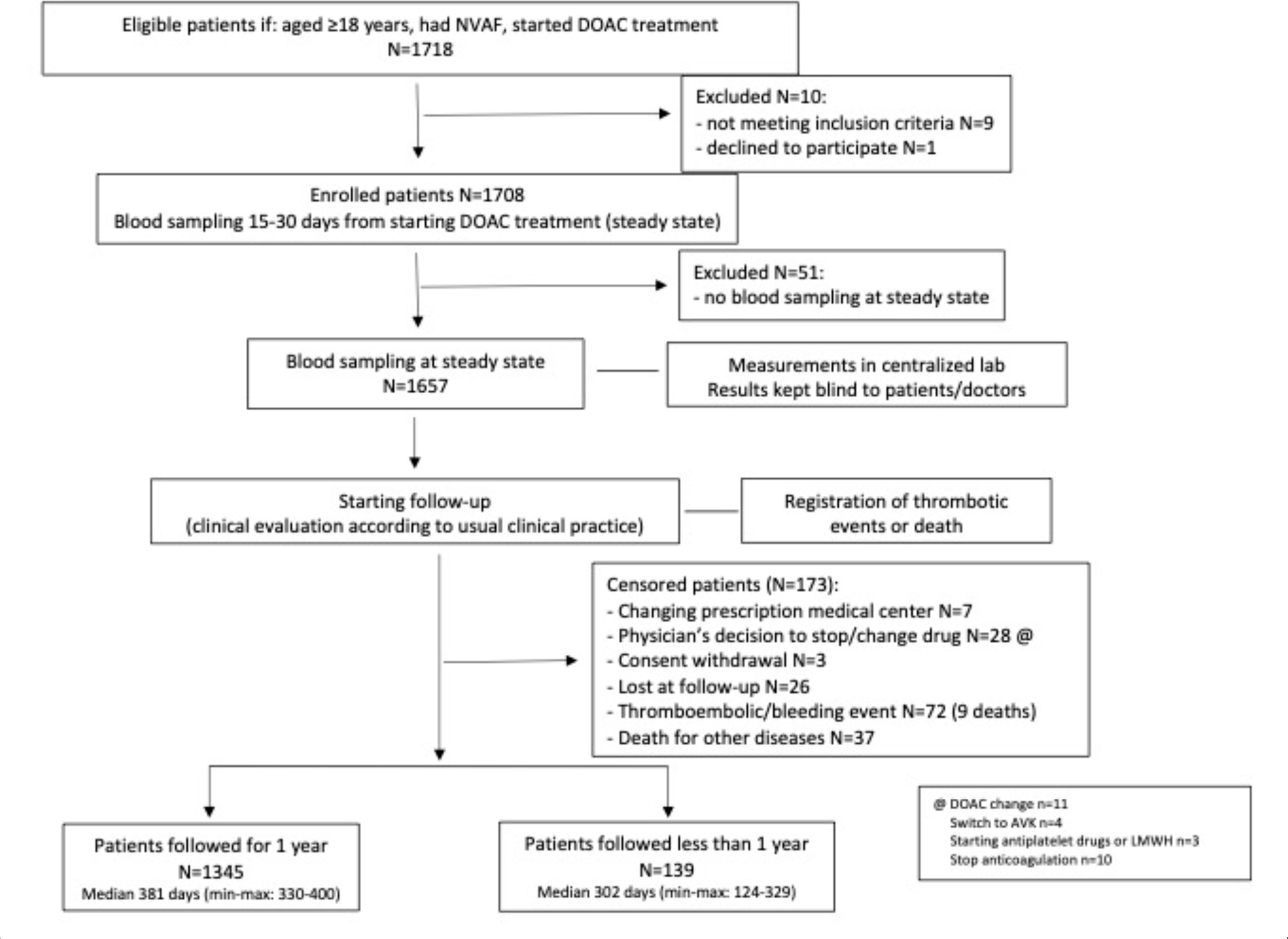
Flowchart of the study

**Table 1.** Baseline characteristics of included patients.

|  |  |
| --- | --- |
| Patients, n | 1657 |
| Participating centers, n | 27 |
| Age, median (min-max), years | 80 (47-100) |
| Males, n (%) | 896 (54.1) |
| BMI, median (min-max) | 26.2 (14.9-68.1) |
| Hemoglobin, median (min-max), (g/dL) | 13.3 (8.0-18.8) |
| Platelets, median (min-max), (<109/L) | 218 (52-700) |
| Creatinine clearance, median (min-max), mL/min | 58.0 (13-246) |
| History of cerebrovascular ischemic disease/ peripheral arterial emboli, n (%) | 186 (11.2) |
| History of cardiovascular disease, n (%) | 284 (17.1) |
| History of gastrointestinal bleeding, n (%) | 22 (1.3) |
| History of cancer, n (%) | 227 (13.7) |
| Hypertension, n (%) | 1472 (88.8) |
| Diabetes, n (%) | 375 (22.6) |
| Liver cirrhosis, n (%) | 14 (0.8) |
| Chronic kidney disease, n (%) | 197 (11.9) |
| Hypothyroidism/Hyperthyroidism, n (%) | 165 (10.0)/62 (3.7) |
| Smokers, n (%) | 190 (11.5) |
| Alcohol intake, n (%) | 58 (3.5) |
| Mental disorders, n (%) | 52 (3.1) |
| Family/social support, n (%) | 1410 (85.1) |
| Drug daily dose, n (%) |  |
| - Apixaban [Standard dose] | 521 (31.4) [336 (65.5)] |
| - Dabigatran [Standard dose] | 221 (13.3) [100 (45.3)] |
| - Edoxaban [Standard dose] | 583 (35.2) [283 (48.6)] |
| - Rivaroxaban [Standard dose] | 332 (20.0) [238 (71.7)] |
| Prescribing accuracy of DOACs<sup>19</sup>: |  |
| - Appropriate, n (%) | 1411 (85.2) |
| Prior AVK treatment, n (%) | 512 (30.9) |
| Use of antiplatelet drugs, n (%) | 382 (23.0) |
| Number of associated drugs, median (min-max) | 3 (0-9) |
| - Antihypertensives, n (%) | 933 (56.3) |
| - Antiarrhythmics, n (%) | 695 (41.9) |
| - Gastroprotectors, n (%) | 655 (39.5) |
| - Antidyslipidemics, n (%) | 585 (35.3) |
| - Thyroid disease drugs, n (%) | 210 (12.7) |
| - Anxiolytics, n (%) | 175 (10.6) |
| - Psychotropics, n (%) | 137 (8.3) |
| - Painkillers, n (%) | 67 (4.0) |
| - Steroids, n (%) | 47 (2.8) |
| - Antiepileptic drugs, n (%) | 29 (1.8) |
| - Nitrates, n (%) | 13 (0.8) |
| - Immunosuppressants, n (%) | 11 (0.7) |
| - Antivirals, n (%) | 7 (0.4) |
| Polytherapy ≥3, n (%) | 1196 (72.2) |
| CHA<sub>2</sub>DS<sub>2</sub>VASc score, median (min-max) | 4 (0-8) |
| CHA<sub>2</sub>DS<sub>2</sub>VASc score ≥4, n (%) | 1072 (64.7) |

BMI: Body-Mass Index; DOAC: direct oral anticoagulants; VKA: vitamin K antagonist

Plasma samples for DOAC measurement were available for all patients at C-trough, and in 1303 patients at C-peak. Results [mean±standard deviation (SD) and min-max] of DOAC levels, at C-trough and at C-peak, are shown in Supplemental Table 1, in addition with the number of plasma samples analyzed for the different DOAC.

### DOAC concentration levels and thrombotic events during follow-up

During a total follow up of 1606 years, thromboembolic outcomes occurred in 21 patients (incidence of 1.31% pt/y): 6 strokes (1 fatal), 1 TIA, 1 VTE, 12 AMI (5 fatal) and 1 superficial vein thrombosis were recorded. Details of patients, who had thrombotic outcomes during follow-up are shown in Supplemental Table 2. Altogether, 46 deaths were recorded (2.8%), 6 of whom were related to thrombotic complications (Supplemental Table 3). DOAC plasma level was the most important independent predictor of occurrence of the primary study outcome according to the best-fitting model (Table 2).

**Table 2.** Effect of standardized plasma DOAC levels on the primary thrombotic outcome endpoint, adjusted for potential confounders.

| Characteristic | First Model (C-trough), n=1657 |  | Second model (C-peak), n=1298 |  |
| --- | --- | --- | --- | --- |
|  | HR | 95% CI | HR | 95% CI |
| Standardized trough DOAC | 0.56 | 0.37 - 0.86 | - | - |
| Standardized peak DOAC | - | - | 0.19 | 0.06-0.66 |
| CHA<sub>2</sub>DS<sub>2</sub>VASc score | 2.01 | 1.02-3.97 | 2.07 | 1.26-3.39 |
| BMI, Kg/m<sup>2</sup> | 0.93 | 0.80-1.08 | 0.95 | 0.82-1.09 |
| Glomerular filtration rate, ml/min | 1.02 | 1.00-1.05 | 1.02 | 0.99-1.05 |
| Low-dose vs Standard dose DOAC | 3.49 | 0.76-16.0 | 2.72 | 0.55-13.5 |
| Antiplatelet treatment (yes vs. no) | 0.28 | 0.03-2.53 | 0.25 | 0.03-1.81 |

Both models were estimated using the Fine & Gray competitive risk regression model. The AIC was 118.4 and 106.1 for the models using C-trough and C-peak, respectively.
BMI: Body-Mass Index; CI: confidence interval; DOAC: Direct oral anticoagulant; HR: hazard ratio

As shown in Figure 2 and Supplemental Table 4, patients with thrombotic outcomes had C- trough DOAC values below the mean value for each drug in 17 (1.6%) cases, whereas 4 (0.66%) cases had values above the mean value. At C-peak, their values were below the mean in13 (1.8%) cases and above in 4 (0.70%) cases. Using standardized C-trough values patients were distributed into classes of increasing levels (Table 3). The highest incidence of thrombotic events (4.82% pt/y) occurred among the 89 patients whose standardized DOAC values were in the lowest class (≤ - 1.00) compared to the mean value; the incidence of events decreased sharply in the other classes. The incidence of events in the lowest class of standardized levels was significantly different than in the sum of all the other classes (4.82% pt/y vs 1.12% pt/y; p= 0.0039). The Kaplan-Meier curves of cumulative thrombotic outcomes occurring in patients in the lowest class of standardized DOAC levels, assessed at C-trough, compared with those in all the higher classes are shown in Figure 3.

**Figure 2.**
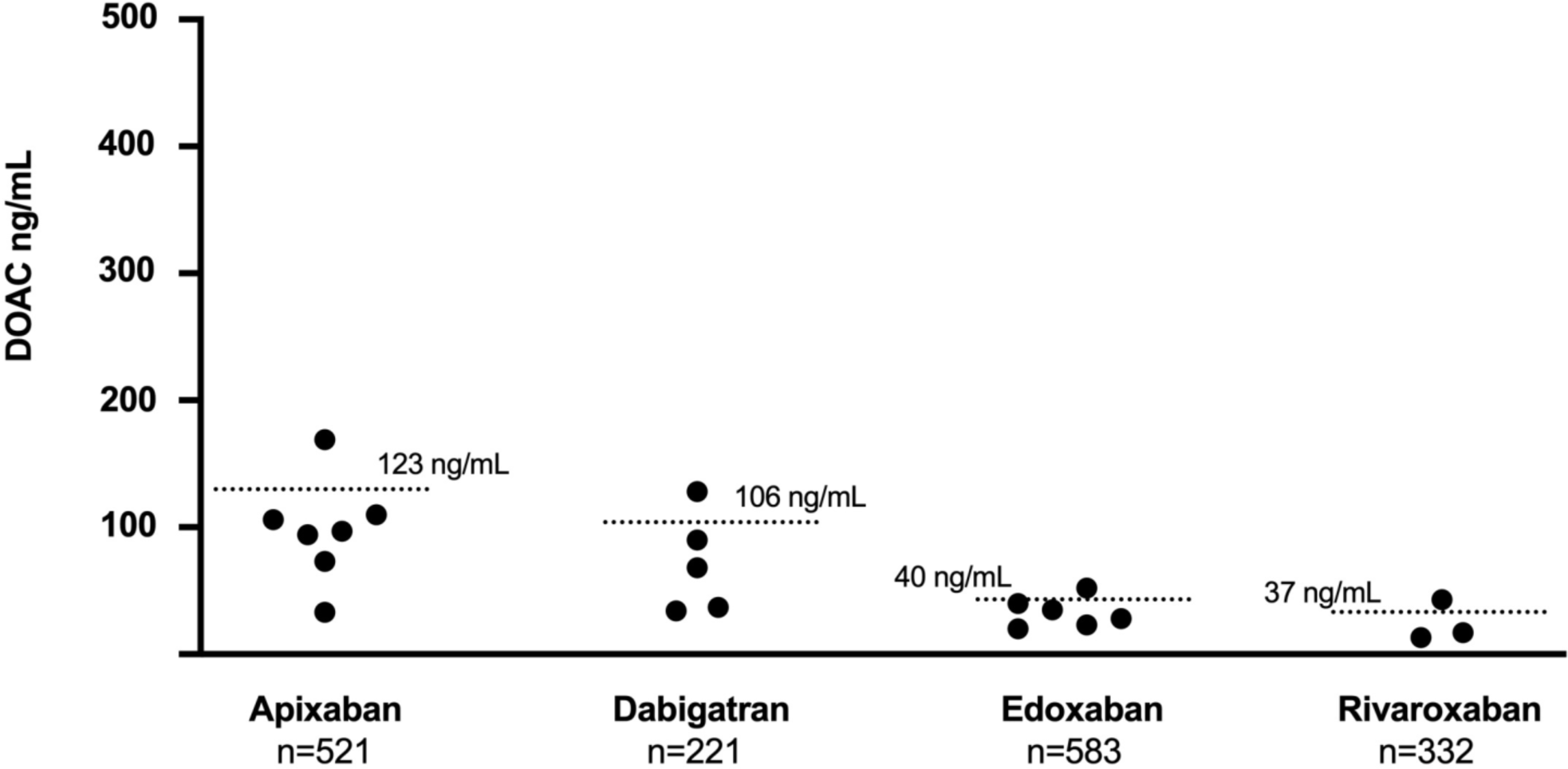
Black dots represent the C-trough DOAC values, assessed at steady state, in atrial fibrillation patients who experienced thrombotic outcomes within the subsequent year of follow-up. Dotted lines represent the mean values of each drug.

**Table 3.**
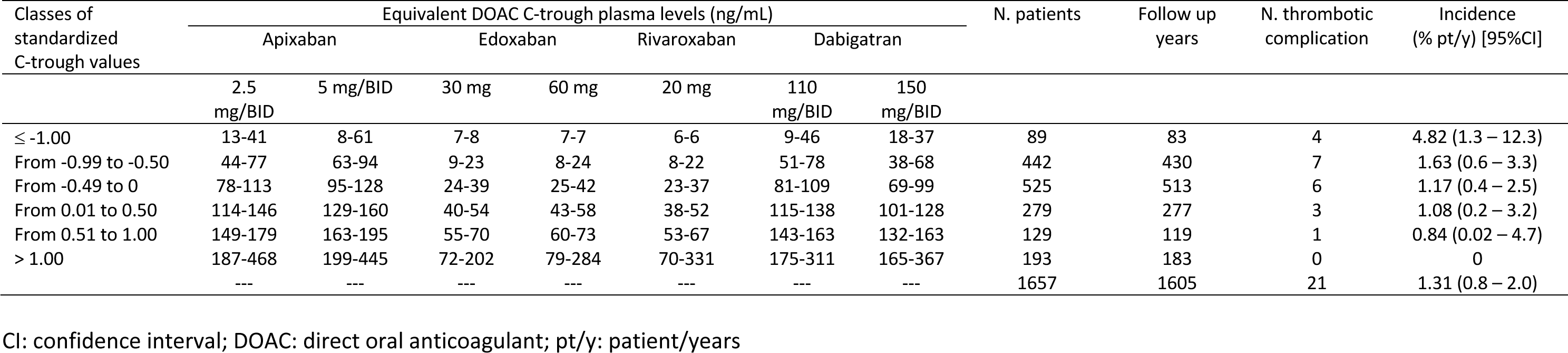
Patient distribution in classes of standardized C-trough values around the mean value (0) for all anticoagulant drugs, with the number of patients and of thrombotic complications recorded in each class. The equivalent DOAC plasma levels at trough for each class are also reported.

**Figure 3.**
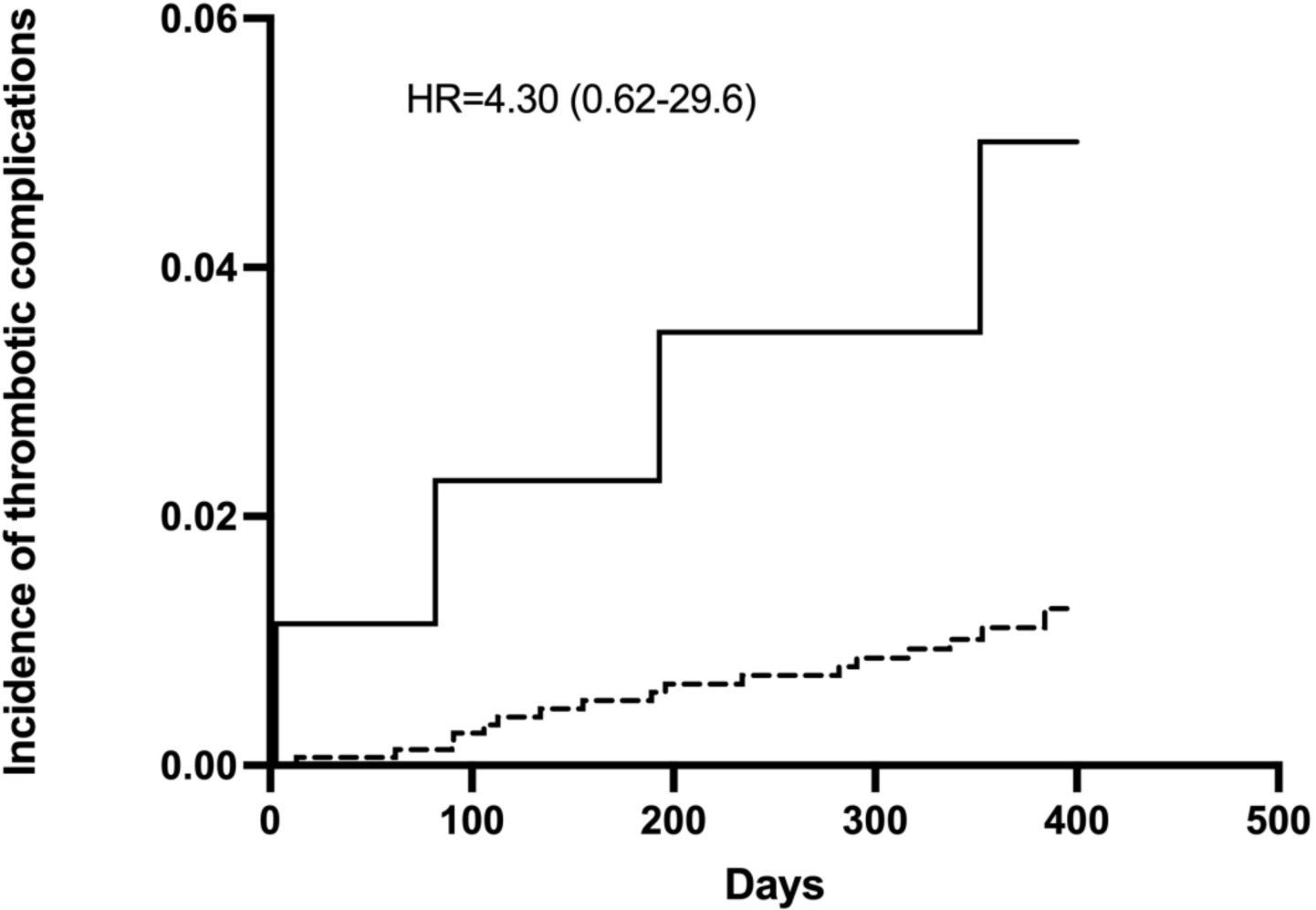
The Kaplan-Meier cumulative event rates for the thrombotic outcomes in patients with DOAC levels in the standardized class σ; - 1.00 (continuous line) and in patients with DOAC levels in the standardized classes > - 1.00 (dotted line) at C-trough.

Based on the present results, we analyzed the hypothetic burden of patients that would be tested using the criterion of CHA2DS2VASc increasing scores, provided that patients have at least one of the following characteristics a) history of ischemic disease (cerebral or cardiovascular), b) have received low-dose DOAC prescription. Table 4 shows the number of patients to be tested, together with the thrombotic events that would (or not) be prevented.

**Table 4.** Analysis of criteria to select patients to be tested; number of patients and thrombotic events that potentially would be avoided (or not) based on our results.

| Criteria to select patients to be tested: | Number of patients who would be tested | Outcomes expected to be avoided | Outcomes that would not be avoided | Number Needed to diagnose |
| --- | --- | --- | --- | --- |
| Patients with increasing CHA<sub>2</sub>DS<sub>2</sub>VASc scores, provided they have at least one of the following conditions: |  |  |  |  |
| a) ischemic disease (cerebral- or cardio-vascular) |  |  |  |  |
| b) on low-dose DOAC treatment |  |  |  |  |
| CHA<sub>2</sub>DS<sub>2</sub>VASc score $\geq 3$ | 892 | 17 | 4 | 223 |
| CHA<sub>2</sub>DS<sub>2</sub>VASc score $\geq 4$ | 767 | 15 | 6 | 128 |
| CHA<sub>2</sub>DS<sub>2</sub>VASc score $\geq 5$ | 472 | 10 | 11 | 43 |
| CHA<sub>2</sub>DS<sub>2</sub>VASc score $\geq 6$ | 168 | 3 | 18 | 9.3 |

The Number needed to diagnose was calculated as = $1 / ((\text{Outcomes Total} / \text{Number of patients}) - (\text{outcomes avoided} / \text{Number of patients}))$
DOAC: direct oral anticoagulant

## DISCUSSION

The MAS Study is the first, observational, multicenter study, which measured DOAC plasma levels in AF patients at the beginning (steady state) of DOAC treatment and prospectively followed-up to record thrombotic complications occurring within one year. DOAC levels were kept blind to patients and to attending physicians and merged with the individual patients only at the end of the study. The main findings of the study are that 17 out of 21 patients who experienced thrombotic outcomes had low plasma activity of the drug at the beginning of treatment; furthermore, the highest incidence of thrombotic complications (4.08% pt/y) occurred in patients whose standardized levels were in the lowest and most distant class from the overall mean.

Efficacy of DOAC treatment for stroke prevention in AF has been largely documented^5–9^ however, a non-negligible incidence of thrombotic events has been recorded in registration trials and clinical practice in patients receiving DOAC.^9–11^ These results pose the question of how to improve clinical management of DOAC-treated AF patients to further reduce the risk of complications. DOAC are administered to AF patients at fixed dose, either standard or low dose in relation to patient characteristics (mainly age and renal function), without the need for laboratory monitoring. Since pharmacological studies, based on pharmacokinetic parameters, have shown a predictable anticoagulant response and the registration trials were conducted at fixed-dose regimens,^1–4,20–24^ it has unanimously been accepted and realized in clinical practice that patients may receive fixed doses, without any need for adapting the dose based on laboratory testing.

However, high inter-individual variability has been confirmed in trials and observational studies,^25,27–29,37–39^ showing that some patients may have low anticoagulant levels and therefore are more exposed to an increased risk of thrombotic complications, especially if at high cardiovascular risk. In a previous study^30^ we observed a relationship between low DOAC levels and thrombotic events, particularly in patients with high CHA2DS2VASc score. Very recently, a monocentric observational study^33^ reported early stroke recurrence in 3% of AF patients with a previous stroke, for whom low plasma levels of apixaban and dabigatran had been detected at steady state.

In the present study, which involved 1657 AF patients treated with the four DOAC currently on the market, we collected data on thrombotic cardiovascular events occurring for a one-year follow-up after blood sampling taken at a steady state to measure the DOAC levels. We found that more than 80% of thrombotic complications occurred in patients with standardized values present in the lowest class, below zero (the overall mean of DOAC levels). Interestingly, both C-peak and C- trough standardized DOAC plasma concentrations had similar predictive capability in our study; however, C-trough measurements are generally easier to obtain.

Though these results seem to indicate that measurement of DOAC levels at a steady state in AF patients may help avoiding most thrombotic complications that may occur during treatment, our results clearly show that it would be unsuitable to measure the drug levels in all patients with AF who start a DOAC treatment to avoid (or reduce) the relatively few thrombotic complications that were recorded during one-year follow-up (21 events, 1.31% pt/y). Nevertheless, it appears clinically relevant to try to improve the efficacy of DOAC treatment by further lowering the incidence of severe thrombotic complications, which would seem achievable by adjusting the daily dose or changing the drug in patients at high thrombotic risk who show low DOAC levels when measured at the beginning of treatment. To this aim, we suggest that different combinations of patient characteristics may be helpful in identifying a criterion which would allow the minimum number of patients to be tested together with the maximum number of thrombotic events that potentially would be avoided.

## Limitations

Our study has limitations. The enrolment was strongly hit by the COVID-19 pandemic, that dramatically blocked many activities at the participating centers. The four available DOAC are not similarly used in our country and, consequently, the number of investigated patients was not equal for each drug. Although rivaroxaban tablets should be taken with food to increase absorption (Product Monograph, Bayer, revision April 2023) in this study blood sampling was performed before the administration of the subsequent DOAC dose, and we are not aware whether patient blood sampling was performed before or after food intake for individual patients, DOAC concentration was measured only once (i.e., 15-30 days after the initiation of treatment).

We therefore cannot exclude possible intra-individual changes in DOAC levels and problems in adherence to treatment during follow-up. However, a previous study reported that the intra-individual variability of DOAC levels was substantially lower than the inter-individual variability.^28^ On the other hand, testing DOAC at different time points during follow up would have been unpractical. However, our study showed an association between very low DOAC levels assessed at the beginning of treatment and occurrence of thrombotic complications in the one-year follow-up, and therefore our results were not influenced by possible DOAC level changes after the time-point of measurement. It is well known that association with potent inducer agents of DOAC catabolism (such as antiepileptic drugs) may significantly reduce DOAC concentration.^40^ The concomitant use of antiepileptic drugs in our study was reported in 28 of the investigated patients, 5 of them had standardized C-trough values in the lowest class around the overall mean (Σ - 1.00), and none of these patients had thrombotic complications during follow-up. All in all, we believe that an accurate analysis of the many factors that may interfere with DOAC concentration would be needed to determine which patients should be tested in future studies. For example, DOAC dosing is still uncertain in some groups of patients, such as those with an extreme body weight. Weight and Body-Mass Index (BMI) are important variables in drug distribution and plasma concentration levels and in individuals with very low or high BMI DOAC measurements may be considered.^19^ In our study population, the number of subjects with BMI <18.5 or >50 (criteria suggested by Steffel et al to define patients with extreme body weight^19^) were small (39 and 1, respectively). The relatively low number of subjects with extreme BMI may have contributed to the non-significant result of BMI as predictor of thrombotic outcomes in the competitive risk score analysis (Table 2).

The strengths of the study are its prospective and multicentric design, the centralization of laboratory tests, which avoided inter-laboratory variations, and the blindness of all test results to patients and treating physicians that avoided possible treatment adjustment during follow-up.

## Future directions

While we are aware that the results of this study are not sufficient to modify current clinical practice, we believe that our findings may pave the way to future studies aimed to definitively assess whether measuring DOAC plasma levels at steady state in selected patients may reduce the incidence of thrombotic complications during follow-up in the setting of AF patients.

## Conclusion

In conclusion, our data show a relationship between low DOAC levels measured at steady state and occurrence of stroke, TIA and VTE, and other thrombotic cardiovascular events in AF patients. The results support the clinical utility of measuring DOAC concentration at the beginning of treatment in special settings of AF patients. Laboratory measurement at steady state may allow to avoid a persistent treatment at insufficient DOAC concentration in patients who are at a very high thrombotic risk. Our results need to be confirmed and expanded with studies in which DOAC treatment is assessed at steady state and adjusted according to the measured levels.

## Funding

The study was promoted and funded by the “Arianna Anticoagulazione” Foundation, which has received an unrestricted grant from the Fondazione “Cassa di Risparmio in Bologna” to support the study. Becton-Dickinson (Franklin Lakes, NJ, USA) provided test tubes for blood sampling, and Diagnostica Stago (Asnieres-sur-Seine, France) provided reagents to perform DOAC.

## Conflict of Interest Disclosures

The authors declare no conflicts of interest related to the present study.

## Data Sharing Statement

For original data, please contact.

## Supplemental material

Table S1-S4

Figure S1

## Non-standard Abbreviations and Acronymis

AF: atrial fibrillation; AIC: Akaike information criteria; AIFA: Italian regulatory agency; AMI: acute myocardial infarction; AVK: anti-vitamin K; BID: twice daily; BMI: Body-Mass Index; CI: confidence interval; CrCl: creatinine clearance; DOAC: direct oral anticoagulants; DVT: deep vein thrombosis; EC: Ethics Committee; FCSA: Italian Federation of Anticoagulation Centers; HR: hazard ratio; pt/y: patient/years; MAS: Measure and See Study; MB: major bleeding; NA: not available; OID: once daily; SD: standard deviation; SVT: superficial vein thrombosis; TIA: transient ischemic attack; VTE: venous thromboembolism

## Data Availability

For original data, please contact

## Clinical centers of the MAS Study group

(in decreasing order of inclusion)

- Sophie Testa, Claudia Dellanoce, Oriana Paoletti, Rossella Morandini, Maurizio Tala. Centro Emostasi e Trombosi, UUOO Laboratorio Analisi chimico-cliniche e microbiologiche, ASST Cremona, Cremona, Italy
- Antonio Ciampa, Martina Gaeta. Centro emostasi, UOC Laboratorio Analisi, Ospedale S.G. Moscati, Avellino, Italy
- Paolo Chiarugi, Monica Casini, Valentina Guerri. UO di Analisi chimico cliniche, Azienda Ospedaliero Universitaria Pisana, Pisa, Italy
- Rita Carlotta Santoro, Piergiorgio Iannaccaro. Centro Emostasi e Trombosi, UO Emofilia e Patologie della Coagulazione, Dipartimento di Ematologia, Oncologia e Medicina Trasfusionale, Azienda Ospedaliero Universitaria Dulbecco, Catanzaro, Italy
- Angela Maria Iannone, Maddalena Campagna. UOSVD Sezione Trasfusionale, Ospedale Don Tonino Bello, Molfetta, Bari, Italy
- Erica De Candia, Maria Adele Alberelli, Maria Basso, Raimondo De Cristofaro, Leonardo Di Gennaro, Antonietta Ferretti, Silvia Sorrentino. UOSD Malattie Emorragiche e Trombotiche, Dipartimento Diagnostica per Immagini, Radioterapia Oncologica ed Ematologia, Fondazione Policlinico Universitario Agostino Gemelli IRCCS, Roma, Italy
- Pasquale Pignatelli, Danilo Menichelli, Daniele Pastori, Mirella Saliola. UOC Medicina Interna e Prevenzione dell’Aterosclerosi, Dipartimento di Medicina Interna e Specialità Mediche, Azienda Ospedaliero-Universitaria Policlinico Umberto I, Roma, Italy
- Elena Maria Faioni, Ilaria Avarello, Cristina Razzari. Servizio Immunologia e Medicina Trasfusionale, Ospedale San Paolo, ASST Santi Paolo e Carlo, Milano, Italy
- Antonio Chistolini, Simona Michela Aprile, Cristina Santoro, Alessandra Serrao. UO Medicina Traslazionale e di Precisione, Dipartimento Medicina Interna e Specialità Mediche, Azienda Ospedaliero-Universitaria Policlinico Umberto I, Roma, Italy
- Maria del Pilar Esteban, Sergio Ricca. UO Laboratorio Analisi, Dipartimento dei Servizi Diagnostici, Ospedale Oglio Po, ASST Cremona, Cremona, Italy
- Marco Marietta, Laura Arletti, Valeria Coluccio, Giulia Debbia, Deborah Grisolia. Struttura Complessa di Ematologia, Policlinico di Modena, Azienda Ospedaliera-Universitaria di Modena, Modena, Italy
- Domizio Serra, Alberto Orselli, Alessandra Pescarollo. Servizio Analisi, Ospedale Evangelico Internazionale, Sede di Castelletto, Genova, Italy
- Sandra Verna, Patrizia Di Gregorio. Servizio di immunoematologia e medicina trasfusionale, Ospedale “SS. Annunziata”, Chieti, Italy
- Giuseppina Cassetti, Mauro Molteni, Mauro Monelli. Medicina Interna, IRCCS Maugeri Milano, Milano, Italy
- Carmelo Paparo, Guido Resani. Laboratorio Analisi, Ospedale Maggiore Chieri, Torino, Italy
- Nicoletta Di Gregorio, Davide Grassi. UOC Medicina Interna e Nefrologia, Presidio Ospedaliero L’Aquila, L’Aquila, Italy
- Corrado Lodigiani, Elena Banfi, Paola Ferrazzi, Luca Librè, Veronica Pacetti, Clara Sacco. Centro Trombosi e Malattie Emorragiche, Humanitas Research Hospital, Rozzano, Milano, Italy
- Paolo Bucciarelli, Ida Martinelli, Maria Abbattista, Andrea Artoni, Marco Capecchi, Francesca Gianniello, Barbara Scimeca. Centro Emofilia e Trombosi Angelo Bianchi Bonomi, Fondazione IRCCS Ca’ Granda Ospedale Maggiore Policlinico, Milano, Italy
- Anna Turrini, Francesca Moretta, Giorgio Parise, Ciro Zeccardo. Laboratorio Analisi Cliniche e Medicina Trasfusionale, IRCCS Ospedale Sacro Cuore Don Calabria, Negrar, Verona, Italy
- Vittorio Fregoni, Massimo Balboni, Federico Leggio. UOC Medicina Generale, Ospedale di Sondalo, Sondalo, Sondrio, Italy
- Daniela Poli, SOD Malattie Aterotrombotiche, Dipartimento Cardiovascolare, Azienda Ospedaliero- Universitaria Careggi, Firenze, Italy
- Luigi Ria, Marina Spagnolo. Centro Emostasi e Trombosi, Medicina Generale e Lungodegenza, Ospedale “Sacro Cuore di Gesù” Gallipoli, Lecce, Italy
- Giovanni Dirienzo, Lavinia Dirienzo, Diana Fuzio. UOSVD Patologia Clinica, Ospedale Della Murgia “Fabio Perinei”, Altamura, Bari, Italy
- Marco Paolo Donadini, Alessandro Squizzato, Walter Ageno, Giovanna Colombo, Silvia Galliazzo, Andrea Gallo, Eleonora Tamborini Permunian, Alexandra Virano. SSD Degenza Breve Internistica, Dipartimento di Medicina Interna, Ospedale di Circolo e Fondazione Macchi, ASST Sette Laghi, Varese, Italy
- Anna Falanga, Luca Barcella, Sara Gamba, Teresa Lerede, Anna Maggioni, Laura Russo, Francesca Schieppati, Federica Zunino. Servizio di Immunoematologia e Medicina Trasfusionale, ASST Papa Giovanni XXIII, Bergamo, Italy
- Giovanni Barillari, Antonella Bertone, Alessandra Poz, Ugo Venturelli. Ambulatorio Malattie Emorragiche e Trombotiche, Medicina Trasfusionale di Udine, Presidio Ospedaliero Universitario “Santa Maria della Misericordia”, Udine, Italy
- Giuseppina Serricchio, Francesca Brevi. UOC Patologia Clinica, Presidio Ospedaliero Sant’Anna, ASST Lariana, San Fermo della Battaglia, Como, Italy

## Notes

### Competing Interest Statement

The authors have declared no competing interest.

### Clinical Trial

NCT03803579

## REFERENCES

1. Connolly SJ, Ezekowitz MD, Yusuf S, Eikelboom J, Oldgren J, Parekh A, Pogue J, Reilly PA, Themeles E, Varrone J, et al. Dabigatran versus warfarin in patients with atrial fibrillation. N Engl J Med. 2009;361:1139–1151.

2. Granger CB, Alexander JH, McMurray JJ, Lopes RD, Hylek EM, Hanna M, Al-Khalidi HR, Ansell J, Atar D, Avezum A, et al. Apixaban versus warfarin in patients with atrial fibrillation. N Engl J Med. 2011;365:981–992. doi: 10.1056/NEJMoa1107039 [doi]

3. Patel MR, Mahaffey KW, Garg J, Pan G, Singer DE, Hacke W, Breithardt G, Halperin JL, Hankey GJ, Piccini JP, et al. Rivaroxaban versus warfarin in nonvalvular atrial fibrillation. N Engl J Med. 2011;365:883–891. doi: 10.1056/NEJMoa1009638[doi]

4. Giugliano RP, Ruff CT, Braunwald E, Murphy SA, Wiviott SD, Halperin JL, Waldo AL, Ezekowitz MD, Weitz JI, Spinar J, et al. Edoxaban versus warfarin in patients with atrial fibrillation. N Engl J Med. 2013;369:2093–2104. doi: 10.1056/NEJMoa1310907

5. Pan KL, Singer DE, Ovbiagele B, Wu YL, Ahmed MA, Lee M. Effects of Non-Vitamin K Antagonist Oral Anticoagulants Versus Warfarin in Patients With Atrial Fibrillation and Valvular Heart Disease: A Systematic Review and Meta-Analysis. J Am Heart Assoc. 2017;6. doi: 10.1161/JAHA.117.005835

6. Durand M, Schnitzer ME, Pang M, Carney G, Eltonsy S, Filion KB, Fisher A, Jun M, Kuo IF, Renoux C, et al. Comparative effectiveness and safety of direct oral anticoagulants versus vitamin K antagonists in nonvalvular atrial fibrillation: a Canadian multicentre observational cohort study. CMAJ Open. 2020;8:E877–E886. doi: 10.9778/cmajo.20200055

7. Lee SR, Choi EK, Kwon S, Jung JH, Han KD, Cha MJ, Oh S, Lip GYH. Effectiveness and Safety of Direct Oral Anticoagulants in Relation to Temporal Changes in Their Use. Circ Cardiovasc Qual Outcomes. 2020;13:e005894. doi: 10.1161/circoutcomes.119.005894

8. Carnicelli AP, Hong H, Connolly SJ, Eikelboom J, Giugliano RP, Morrow DA, Patel MR, Wallentin L, Alexander JH, Cecilia Bahit M, et al. Direct Oral Anticoagulants Versus Warfarin in Patients With Atrial Fibrillation: Patient-Level Network Meta-Analyses of Randomized Clinical Trials With Interaction Testing by Age and Sex. Circulation. 2022;145:242–255. doi: 10.1161/CIRCULATIONAHA.121.056355

9. Lip GYH, Keshishian A, Li X, Hamilton M, Masseria C, Gupta K, Luo X, Mardekian J, Friend K, Nadkarni A, et al. Effectiveness and Safety of Oral Anticoagulants Among Nonvalvular Atrial Fibrillation Patients. Stroke. 2018;49:2933–2944. doi: 10.1161/strokeaha.118.020232

10. Poli D, Antonucci E, Ageno W, Bertu L, Migliaccio L, Martinese L, Pilato G, Testa S, Palareti G. Oral anticoagulation in very elderly patients with atrial fibrillation: Results from the prospective multicenter START2-REGISTER study. PLoS One. 2019;14:e0216831. doi: 10.1371/journal.pone.0216831

11. Harrison SL, Buckley BJR, Ritchie LA, Proietti R, Underhill P, Lane DA, Lip GYH. Oral anticoagulants and outcomes in adults ≥80 years with atrial fibrillation: A global federated health network analysis. J Am Geriatr Soc. 2022;70:2386–2392. doi: 10.1111/jgs.17884

12. Reilly PA, Lehr T, Haertter S, Connolly SJ, Yusuf S, Eikelboom JW, Ezekowitz MD, Nehmiz G, Wang S, Wallentin L, et al. The effect of dabigatran plasma concentrations and patient characteristics on the frequency of ischemic stroke and major bleeding in atrial fibrillation patients: the RE-LY Trial (Randomized Evaluation of Long-Term Anticoagulation Therapy). J Am Coll Cardiol. 2014;63:321–328. doi: 10.1016/j.jacc.2013.07.104

13. Girgis IG, Patel MR, Peters GR, Moore KT, Mahaffey KW, Nessel CC, Halperin JL, Califf RM, Fox KA, Becker RC. Population pharmacokinetics and pharmacodynamics of rivaroxaban in patients with non-valvular atrial fibrillation: results from ROCKET AF. J Clin Pharmacol. 2014;54:917–927. doi: 10.1002/jcph.288

14. Zeitouni M, Giczewska A, Lopes RD, Wojdyla DM, Christersson C, Siegbahn A, De Caterina R, Steg PG, Granger CB, Wallentin L, et al. Clinical and Pharmacological Effects of Apixaban Dose Adjustment in the ARISTOTLE Trial. J Am Coll Cardiol. 2020;75:1145–1155. doi: 10.1016/j.jacc.2019.12.060

15. Salazar DE, Mendell J, Kastrissios H, Green M, Carrothers TJ, Song S, Patel I, Bocanegra TS, Antman EM, Giugliano RP, et al. Modelling and simulation of edoxaban exposure and response relationships in patients with atrial fibrillation. Thromb Haemost. 2012;107:925–936. doi: 11-08- 0566 [pii] 10.1160/TH11-08-0566 [doi]

16. Ruff CT, Giugliano RP, Braunwald E, Morrow DA, Murphy SA, Kuder JF, Deenadayalu N, Jarolim P, Betcher J, Shi M, et al. Association between edoxaban dose, concentration, anti-Factor Xa activity, and outcomes: an analysis of data from the randomised, double-blind ENGAGE AF-TIMI 48 trial. Lancet. 2015;385:2288–2295. doi: 10.1016/s0140-6736(14)61943-7

17. Tripodi A. To measure or not to measure direct oral anticoagulants before surgery or invasive procedures. J Thromb Haemost. 2016;14:1325–1327. doi: 10.1111/jth.13344

18. Douxfils J, Adcock DM, Bates SM, Favaloro EJ, Gouin-Thibault I, Guillermo C, Kawai Y, Lindhoff- Last E, Kitchen S, Gosselin RC. 2021 Update of the International Council for Standardization in Haematology Recommendations for Laboratory Measurement of Direct Oral Anticoagulants. Thromb Haemost. 2021;121:1008–1020. doi: 10.1055/a-1450-8178

19. Steffel J, Collins R, Antz M, Cornu P, Desteghe L, Haeusler KG, Oldgren J, Reinecke H, Roldan- Schilling V, Rowell N, et al. 2021 European Heart Rhythm Association Practical Guide on the Use of Non-Vitamin K Antagonist Oral Anticoagulants in Patients with Atrial Fibrillation. Europace. 2021;23:1612–1676. doi: 10.1093/europace/euab065

20. Kubitza D, Becka M, Voith B, Zuehlsdorf M, Wensing G. Safety, pharmacodynamics, and pharmacokinetics of single doses of BAY 59-7939, an oral, direct factor Xa inhibitor. Clin Pharmacol Ther. 2005;78:412–421. doi: S0009-9236(05)00281-X [pii] 10.1016/j.clpt.2005.06.011

21. Stangier J, Rathgen K, Staehle H, Gansser D, Roth W. The pharmacokinetics, pharmacodynamics and tolerability of dabigatran etexilate, a new oral direct thrombin inhibitor, in healthy male subjects. Brit J Clin Pharmacol. 2007;64:292–303.

22. Eriksson BI, Quinlan DJ, Weitz JI. Comparative pharmacodynamics and pharmacokinetics of oral direct thrombin and factor xa inhibitors in development. Clin Pharmacokinet. 2009;48:1–22.

23. Frost C, Nepal S, Wang J, Schuster A, Byon W, Boyd RA, Yu Z, Shenker A, Barrett YC, Mosqueda- Garcia R, et al. Safety, pharmacokinetics and pharmacodynamics of multiple oral doses of apixaban, a factor Xa inhibitor, in healthy subjects. Br J Clin Pharmacol. 2013;76:776–786. doi: 10.1111/bcp.12106 [doi]

24. Parasrampuria DA, Truitt KE. Pharmacokinetics and Pharmacodynamics of Edoxaban, a Non-Vitamin K Antagonist Oral Anticoagulant that Inhibits Clotting Factor Xa. Clin Pharmacokinet. 2016;55:641–655. doi: 10.1007/s40262-015-0342-7

25. Testa S, Tripodi A, Legnani C, Pengo V, Abbate R, Dellanoce C, Carraro P, Salomone L, Paniccia R, Paoletti O, et al. Plasma levels of direct oral anticoagulants in real life patients with atrial fibrillation: Results observed in four anticoagulation clinics. Thromb Res. 2016;137:178–183. doi: 10.1016/j.thromres.2015.12.001

26. Testa S, Dellanoce C, Paoletti O, Cancellieri E, Morandini R, Tala M, Zambelli S, Legnani C. Edoxaban plasma levels in patients with non-valvular atrial fibrillation: Inter and intra-individual variability, correlation with coagulation screening test and renal function. Thromb Res. 2019;175:61–67. doi: 10.1016/j.thromres.2019.01.008

27. Kampouraki E, Avery P, Biss T, Wynne H, Kamali F. Assessment of exposure to direct oral anticoagulants in elderly hospitalised patients. Br J Haematol. 2021;195:790–801. doi: 10.1111/bjh.17899

28. Toorop MMA, van Rein N, Nierman MC, Vermaas HW, Huisman MV, van der Meer FJM, Cannegieter SC, Lijfering WM. Inter- and intra-individual concentrations of direct oral anticoagulants: The KIDOAC study. J Thromb Haemost. 2022;20:92–103. doi: 10.1111/jth.15563

29. Edwina AE, Dia N, Dreesen E, Vanassche T, Verhamme P, Spriet I, Van der Linden L, Tournoy J. Insights into the Pharmacokinetics and Pharmacodynamics of Direct Oral Anticoagulants in Older Adults with Atrial Fibrillation: A Structured Narrative Review. Clin Pharmacokinet. 2023;62:351–373. doi: 10.1007/s40262-023-01222-w

30. Testa S, Paoletti O, Legnani C, Dellanoce C, Antonucci E, Cosmi B, Pengo V, Poli D, Morandini R, Testa R, et al. Low drug levels and thrombotic complications in high-risk atrial fibrillation patients treated with direct oral anticoagulants. J Thromb Haemost. 2018;16:842–848. doi: 10.1111/jth.14001

31. Testa S, Legnani C, Antonucci E, Paoletti O, Dellanoce C, Cosmi B, Pengo V, Poli D, Morandini R, Testa R, et al. Drug levels and bleeding complications in atrial fibrillation patients treated with direct oral anticoagulants. J Thromb Haemost. 2019;17:1064–1072. doi: 10.1111/jth.14457

32. Rizos T, Meid AD, Huppertz A, Dumschat C, Purrucker J, Foerster KI, Burhenne J, Czock D, Jenetzky E, Ringleb PA, et al. Low Exposure to Direct Oral Anticoagulants Is Associated with Ischemic Stroke and Its Severity. J Stroke. 2022;24:88–97. doi: 10.5853/jos.2020.04952

33. Siedler G, Macha K, Stoll S, Plechschmidt J, Wang R, Gerner ST, Strasser E, Schwab S, Kallmunzer B. Monitoring of direct oral anticoagulants plasma levels for secondary stroke prevention. J Thromb Haemost. 2022;20:1138–1145. doi: 10.1111/jth.15677

34. Nosáľ V, Petrovičová A, Škorňová I, Bolek T, Dluhá J, Stančiaková L, Sivák Š, Babálová L, Hajaš G, Staško J, et al. Plasma levels of direct oral anticoagulants in atrial fibrillation patients at the time of embolic stroke: a pilot prospective multicenter study. Eur J Clin Pharmacol. 2022;78:557–564. doi: 10.1007/s00228-022-03280-8

35. Gosselin RC, Marlar RA. Preanalytical Variables in Coagulation Testing: Setting the Stage for Accurate Results. Semin Thromb Hemost. 2019;45:433–448. doi: 10.1055/s-0039-1692700

36. Fine JPG, R.J. A proportional hazards model for the subdistributionof a competing risk. J Am Stat Assoc. 1999;94:496–509.

37. Gouin-Thibault I, Delavenne X, Blanchard A, Siguret V, Salem JE, Narjoz C, Gaussem P, Beaune P, Funck-Brentano C, Azizi M, et al. Interindividual variability in dabigatran and rivaroxaban exposure: contribution of ABCB1 genetic polymorphisms and interaction with clarithromycin. J Thromb Haemost. 2017;15:273–283. doi: 10.1111/jth.13577

38. Siguret V, Abdoul J, Delavenne X, Curis E, Carlo A, Blanchard A, Salem JE, Gaussem P, Funck-Brentano C, Azizi M, et al. Rivaroxaban pharmacodynamics in healthy volunteers evaluated with thrombin generation and the active protein C system: Modeling and assessing interindividual variability. J Thromb Haemost. 2019;17:1670–1682. doi: 10.1111/jth.14541

39. Foulon-Pinto G, Lafuente C, Jourdi G, Le Guen J, Tall F, Puymirat E, Delrue M, Riviere L, Ketz F, Gouin-Thibault I, et al. Assessment of DOAC in GEriatrics (ADAGE study): rivaroxaban/apixaban concentrations and thrombin generation profiles in NVAF very elderly patients. Thromb Haemost. 2023;123:402–414. doi: 10.1055/a-1981-1763

40. Sennesael AL, Larock AS, Hainaut P, Lessire S, Hardy M, Douxfils J, Spinewine A, Mullier F. The Impact of Strong Inducers on Direct Oral Anticoagulant Levels. Am J Med. 2021;134:1295–1299. doi: 10.1016/j.amjmed.2021.06.003

